# Natural and hybrid immunity following four COVID-19 waves in a South African cohort

**DOI:** 10.1101/2022.06.20.22276647

**Authors:** Heather J Zar, Rae MacGinty, Lesley Workman, Maresa Botha, Marina Johnson, Adam Hunt, Tiffany Bird, Mark P Nicol, Stefan Flasche, Billy J Quilty, David Goldblatt

**Author notes:** **Correspondence:** Prof David Goldblatt, Great Ormond Street Institute of Child Health Biomedical Research Centre, University College London & Great Ormond Street Children’s Hospital NHS Foundation Trust, London, UK.

## Abstract

**Background:** More than half the global population has been exposed to SARS-CoV-2. Naturally induced immunity influences the outcome of subsequent exposure to variants and vaccine responses. We measured anti-spike IgG responses to explore the basis for this enhanced immunity.

**Methods:** A prospective cohort study in a South African community through the ancestral/beta/delta/omicron SARS-CoV-2 waves. Health seeking behaviour/illness were recorded and post-wave serum samples probed for IgG to Spike (CoV2-S-IgG). To estimate protective CoV2-S-IgG threshold levels, logistic functions were fit to describe the correlation of CoV2-S-IgG measured before a wave and the probability for seroconversion/boosting thereafter for unvaccinated and vaccinated adults.

**Findings:** Despite little disease, 176/339 (51·9%) participants were seropositive following wave 1, rising to 74%, 89·8% and 97·3% after waves 2, 3 and 4 respectively. CoV2-S-IgG induced by natural exposure protected against subsequent SARS-CoV-2 infection with the greatest protection for beta and the least for omicron. Vaccination induced higher CoV2-S-IgG in seropositive compared to naïve vaccinees. Amongst seropositive participants, proportions above the 50% protection against infection threshold were 69% (95% CrI: 62, 72) following 1 vaccine dose, 63% (95% CrI: 63, 75) following 2 doses and only 11% (95% CrI: 7, 14) in unvaccinated during the omicron wave.

**Interpretation:** Naturally induced CoV2-S-IgG do not achieve high enough levels to prevent omicron infection in most exposed individuals but are substantially boosted by vaccination leading to significant protection. A single vaccination in those with prior immunity is more immunogenic than 2 doses in a naïve vaccinee and thus may provide adequate protection.

**Funding:** UK NIH GECO award (GEC111), Wellcome Trust Centre for Infectious Disease Research in Africa (CIDRI), Bill & Melinda Gates Foundation, USA (OPP1017641, OPP1017579) and NIH H3 Africa (U54HG009824, U01AI110466]. HZ is supported by the SA-MRC. MPN is supported by an Australian National Health and Medical Research Council Investigator Grant (APP1174455). BJQ is supported by a grant from the Bill and Melinda Gates Foundation (OPP1139859). Stefan Flasche is supported by a Sir Henry Dale Fellowship jointly funded by the Wellcome Trust and the Royal Society (Grant number 208812/Z/17/Z).

**Research in context:** 

**Evidence before this study:** Natural infection with ancestral SARS-CoV-2 virus provides partial protection against re-infection with the same and closely related SARS-CoV-2 variants, but higher rates of re-infection have been described with Omicron. In addition, vaccination against SARS-CoV2 provides relatively lower protection against symptomatic Omicron infection than for other variants. Hybrid immunity, a combination of immunity induced by natural infection and vaccination is of critical interest due to the high incidence of natural infection in many populations and increased availability of vaccination. Vaccination following infection may provide more robust immunity than either infection or vaccination alone, but there are limited data on the impact of hybrid immunity for protection against different variants or on the optimal vaccination strategy following natural infection.

**Added value of this study:** We leveraged a unique South African birth cohort in a poor peri-urban area, to longitudinally investigate infection, illness and serological responses to natural exposure to SARS-CoV-2 over 4 waves of the pandemic in healthy mothers. We also investigated the impact of prior natural exposure on BNT162b2 mRNA vaccine responses. We used this information to derive estimates of levels of spike-specific IgG associated with protection for subsequent infection following natural or hybrid immunity. Despite little disease, most participants were seropositive with rates rising from 52% to 74%, 90% and 97% after waves 1, 2, 3 and 4 respectively. Antibodies to spike protein induced by natural exposure protected against subsequent infection with the greatest protection for beta and the least for omicron. Antibody levels following vaccination were significantly higher in those who were seropositive prior to vaccine, compared to those seronegative. Amongst seropositive participants, proportions above the 50% protection against infection threshold were 69% following 1 vaccine dose, 63% following 2 doses and only 11% in unvaccinated during the omicron wave. In those seropositive prior to vaccination no significant increase in antibody levels occurred after the 2^nd^ dose of vaccine, unlike the increase in seronegative participants. A single dose of vaccine in seropositive individuals induced higher antibody concentrations than two doses in seronegative recipients.

**Implications of all the available evidence:** Naturally induced spike antibodies do not achieve high enough levels to prevent omicron infection in most exposed individuals but are substantially boosted by vaccination leading to significant protection. A single vaccination in those with prior natural immunity is more immunogenic than 2 doses in seronegative people and may provide adequate protection against omicron and other variants. Vaccination programs in populations with high seroprevalence using a single vaccination as a primary strategy should be considered.

## Background

Since the start of the pandemic in late 2019, it is estimated that there have been approximately 520 million confirmed cases of COVID-19, including more than 6·2 million deaths (https://covid19.who.int). Many SARS-CoV-2 infections are asymptomatic and the latest global estimates suggest that half the human population are seropositive as a consequence of exposure.^1^ While natural infection with ancestral SARS-CoV-2 virus provides partial protection against re-infection with the same and closely related SARS-CoV-2 variants,^2,3^ infection with Omicron, antigenically the most distant of the variants of concern to the ancestral wild type strain,^4^ has been associated with higher rates of re-infection.^5,6^ Natural infection with SARS-CoV-2 induces both humoral and cellular immunity and protection against re-infection is likely to be the result of a combination of receptor binding domain antibodies preventing SARS-CoV-2 interaction with ACE2 receptor, thus preventing infection and T cells, specific for a variety of antigens, stopping or modulating the progression to symptomatic and or serious disease and death. Primary immunisation with existing spike-containing authorised vaccines has provided relatively poor protection against symptomatic Omicron infection, most likely due to the variants escape from vaccine-induced immunity secondary to critical mutations in the Receptor Binding Domain.^7^

While both binding and neutralising antibody are recognised as correlates of protection against SARS-CoV-2 infection,^8,9^ much of the focus on antibody correlates has been in relation to vaccine induced immunity,^10,11^ with a focus on future vaccine licensure.^12^ Relatively little is understood of natural immunity and the relationship between antibodies induced after exposure to SARS-CoV-2 and subsequent protection from infection. However such immunity, induced after natural infection, is becoming of critical interest due to the observation that vaccination following infection may provide more robust immunity than either infection or vaccination alone.^13-16^ This so called hybrid immunity^17^ is associated with a breadth of variant recognition that appears to be a consequence of immune maturation.^18^

South Africa has experienced four well-defined SARS-CoV-2 waves of infection; the first driven by the ancestral (Wuhan) strain, the second dominated (>95%) by beta-variant (B.1.351),^2^ the third predominantly due to the delta-variant and the 4^th^ wave due to the Omicron variant.^19^ We leveraged a unique South African birth cohort in a poor peri-urban area, to longitudinally investigate infection, illness and serological responses to natural exposure to SARS-CoV-2 in mothers over 4 waves of the pandemic as well as to study responses to SARS-CoV-2 vaccine to investigate the impact of previous natural exposure on vaccine responses. We used this information to derive estimates of levels of spike-specific IgG associated with protection from subsequent infection following natural or hybrid immunity.

## Methods

We studied participants in an established South African birth cohort, the Drakenstein Child Health Study (DCHS),^20^ using a convenience sample of maternal participants through the COVID-19 pandemic from 6 March 2020 to 28 February 2022, spanning four waves. The convenience sample included sequential mothers attending follow-up visits with their children with blood sampling through all 4 waves of the pandemic. The study is situated in a low-income peri-urban community, in which there is a strong primary health care program, well established study surveillance systems for illness and high cohort retention as previously described.^20^ Illness and health seeking behaviour were monitored throughout and additional study visits through each wave were initiated with serum samples obtained.

Serological responses to SARS-CoV2 were measured in 4 matched sera obtained following each of the 4 waves. These were defined by the SA National Institute of Communicable Diseases as wave 1 (ancestral strain) week 24-35 2020, wave 2 (beta variant) week 48 2020-week 5 2021, wave 3 (delta variant) week 19-37 2021 and wave 4 (omicron variant) week 45 2021-week 3 2022.^21^ A national program for SARS-CoV2 vaccination began for health care workers from March 2021 providing a single dose of Ad.26COV2.S (Johnson & Johnson vaccine; AD26.COV.2.S); this was broadened to include all adults (>18 years) from June 2021, in which a single dose Ad26.COV.2.S or 2 doses of BNT162b2 (Pfizer-BioNTech) vaccine (given 6 weeks apart) became available. Booster doses of either AD26.COV.2.S or BNT162b2 became available from January 2022. The national program is the only source of SARS-CoV-2 vaccination available in South Africa.

The study was approved by the Human Research Ethics Committee, Faculty of Health Sciences University of Cape Town. Mothers provided written informed consent which was renewed annually.

### Antibody measurements

Serum samples from mothers were tested for IgG to spike (S) protein derived from ancestral SARS-CoV-2 (S-ancestral), beta (S-beta), delta (S-delta) or Omicron (S-omicron) variants using the Meso Scale discovery platform (MSD® Rockville, MD) as described before^22^. The detection of S-ancestral IgG in this assay is highly sensitive and specific for exposure to SARS-CoV-2 and hence was used to define seropositivity (S-ancestral ≥1·09 WHO BAU/ml). Geometric mean concentrations (95% CI) of IgG levels (GMC) for SARS-CoV2 antibodies were calculated. IgG to spike from different strains cross-reacts but higher titres are generated to the infecting strain therefore a ratio of variant S-IgG: S-ancestral IgG was calculated.

### Statistical analysis

Data were analyzed using STATA 14.1 (STATA Corporation, College Station, TX USA) and R (R core team 2021, version 4.1.2). Data were summarised as frequencies (percent) if categorical and median (interquartile range (IQR)) if continuous. Wilcoxon rank-sum test (Mann-Whitney U test), Wilcoxon signed-rank test and Chi-square or Fisher’s exact were used for crude comparisons, as appropriate. Seropositivity was measured longitudinally though each wave; once vaccinated, a participant was excluded from calculation of seroprevalence. A Kaplan-Meier plot was used to calculate the time in which unvaccinated participants became seropositive through the 4 waves; a participant was censored at the time of seropositivity.

Generalised estimating equations (GEE) were used to identify risk factors associated with seropositivity over the waves. A binomial distribution and logit link function, as well as robust standard errors to account for the presence of heteroscedascity, were used in generating the GEE models. The model was adjusted for age, HIV infection, marital status, maternal education, maternal employment, household income, household size, maternal smoking, asthma diagnosis and maternal weight.

To estimate threshold levels of antibodies induced by prior exposure or vaccine which may protect against subsequent SARS-CoV-2 infection, logistic functions were fit to spike IgG titres measured before and after the beta, delta and omicron waves. The software package R2Jags was used for Bayesian model fitting. Similar to a logistic regression the probability of seroconversion (defined as titres increasing by more than 1% post wave) after the beta, delta and omicron waves was estimated as a function of the amount of the antibody prior to a wave, but using a more flexible link function. This allowed estimation of infection attack rates in naïve or vaccinated individuals, the maximal protection achievable from naturally derived or vaccine induced antibodies and antibody thresholds associated with specific levels of protection. The model code is available from the github repository: https://github.com/bquilty25/covid_seroconv.

## Results

The detailed characteristics of 339 mothers [median age 32·9y (IQR 28·9; 37·2y)] participating in this study are summarised in Table S1. Participants were predominantly of low socioeconomic status and self-reported maternal smoking occurred in 124 (36·6%).

There were 69 (20·4%) HIV-infected mothers, all were well established on antiretroviral therapy (ART) for a median (IQR) of 8·4 (7·5,11·0) years. The median household size was 5 (4-6) people. During the study period there were 18 (5·3%) PCR-confirmed SARS-CoV-2 infections, 3 COVID-related hospitalizations and no deaths. Median (IQR) follow-up over this period was 495 (475; 517) days with a median of 184 days between blood sampling following wave 1 and 2 and 157 days and 132 days between samples for the two subsequent waves. Two (0·6%) mothers were vaccinated with AD26.COV.2.S before their 2^nd^ wave sample, 95 (28·0%) were vaccinated before their 3^rd^ wave sample (63 with BNT162b2 dose 1; 13 with 2 BNT162b2 doses and 19 with a dose of AD26.COV.2.S) and 154 (45·4%) were vaccinated before the 4^th^ wave (127 with at least 1 BNT162b2 dose, 66 with 2 BNT162b2 doses and 27 with a single dose of AD26.COV.2.S) (Figure S1).

Despite little COVID illness, 176 (51·9%) mothers were seropositive following wave 1. Amongst unvaccinated mothers levels of seropositivity increased to 74·3% (250/337) after wave 2, 89·8% (219/244) after wave 3 and 97·3% (180/185) after wave 4 (Table 1). Only 5 unvaccinated mothers (2·7%) remained seronegative throughout all four waves. Multivariate analysis of factors associated with seropositivity indicated that, age, HIV infection and maternal weight were positively associated with seropositivity across the 4 waves in the unadjusted analysis, while current cigarette smoking was inversely associated (Table S2). In the adjusted model, those in more crowded households had greater odds of seropositivity over the 4 waves (adjusted OR=1·14, 95% CI: 1·02; 1·27) and current smoking was associated with seronegativity (adjusted OR=0·43, 95% CI: 0·28; 0·66) but none of the other covariates remained significant in the adjusted model.

**Table 1.**
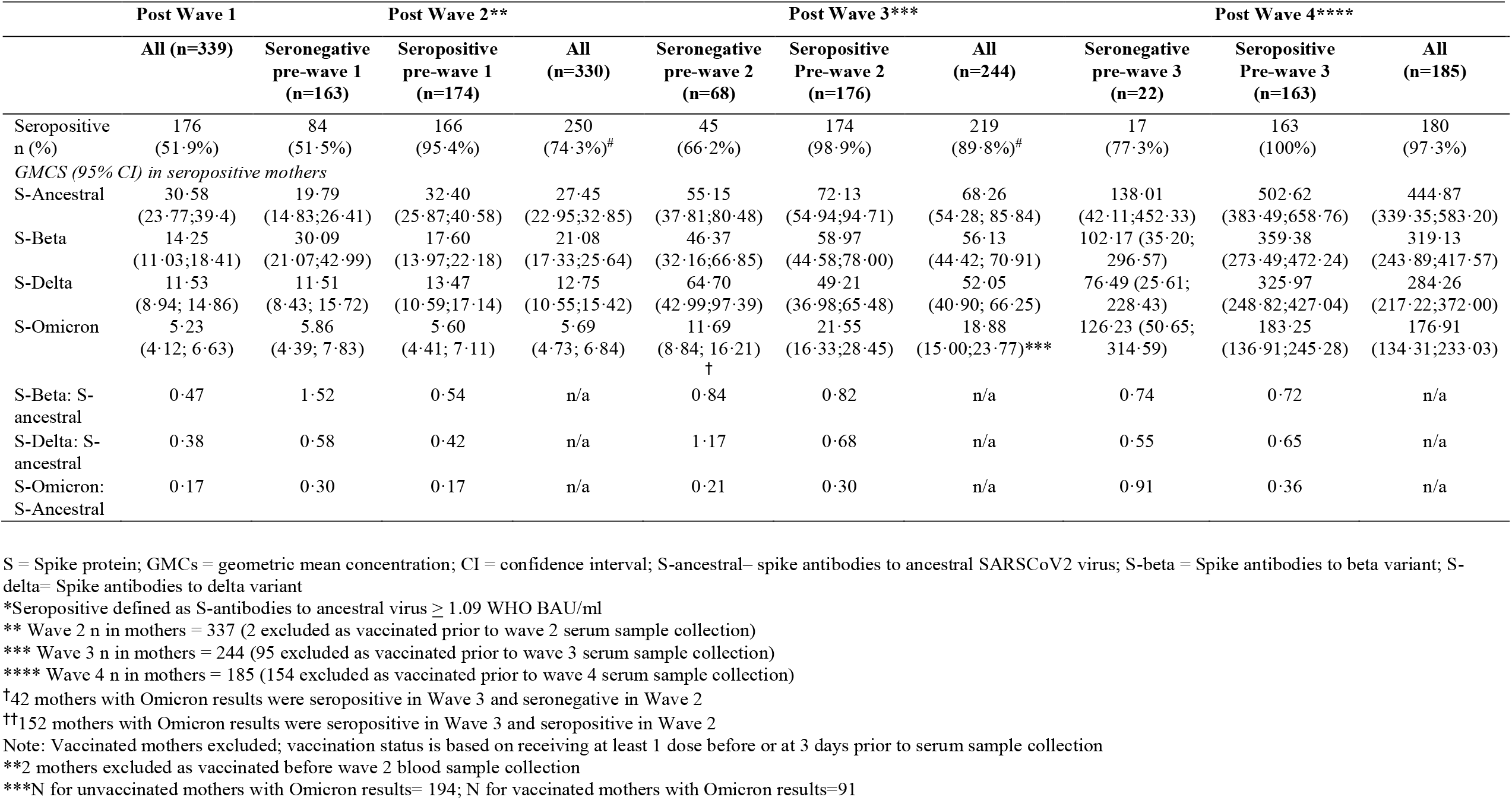
Anti-spike IgG concentrations (GMC, 95%CI) in unvaccinated seropositive mothers following each wave of SARS-CoV-2. Mothers are stratified by their serostatus prior to the wave.

While 52% of seronegative mothers seroconverted following the Wuhan and Beta variant waves (1 and 2), 66% and 77% seroconverted after exposure to the Delta (wave 3) and the Omicron strain (wave 4) respectively consistent with greater transmissibility of these variants of concern (Table 1). Amongst unvaccinated mothers, the highest anti-spike IgG concentrations for all variants were seen after the Omicron wave and concentrations were significantly higher after each wave for mothers seropositive prior to the wave compared to seronegatives consistent with natural priming (Table 1, fig 1A, 1B). Furthermore seronegative mothers demonstrated a ratio of variant to wild-type Spike IgG of >1·0 indicating a primary response to the variant of concern (VOC) dominant during the wave while seropositive mothers, despite an increase in S-IgG, had VOC:WT ratios <1 indicating possible imprinting following the original exposure. A small number of mothers reverted to become seronegative following waves 2 and 3 [8 (2·4%) and 2 (0·8%) respectively] but none following wave 4.

**Figure 1.**
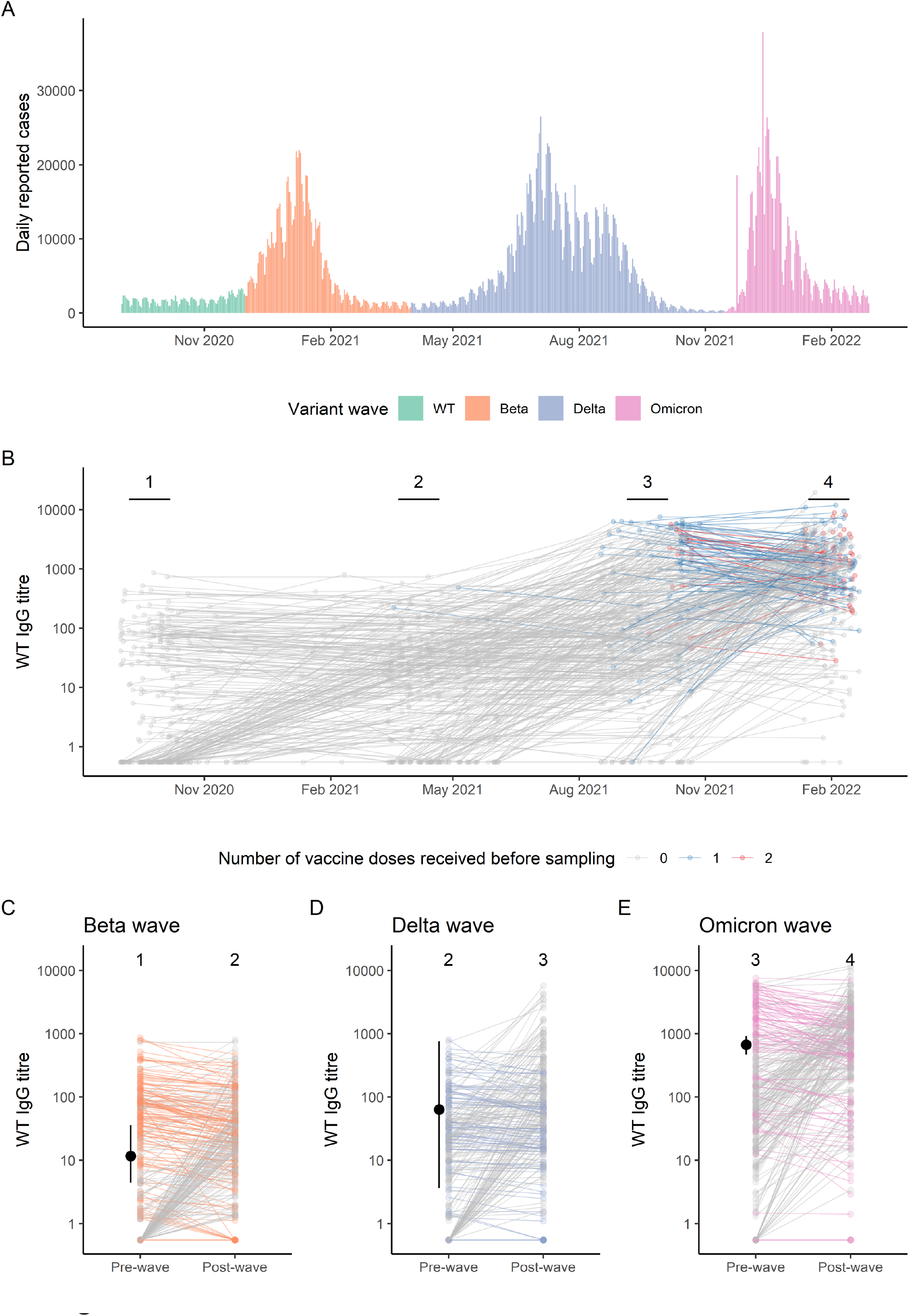
Progression of serostatus during the course of the Covid-19 pandemic in South Africa, and estimated thresholds of protection against seroconversion. A. Daily reported cases in South Africa from September 2020 to March 2022, coloured by predominant circulating serotype, from https://covid19.who.int/WHO-COVID-19-global-data.csv. B. Individual level S-ancestral (WT) IgG titres over time, coloured by vaccine status prior to sampling. C. Wave specific change in S-ancestral (WT) IgG titres over the course of the beta, delta, and omicron waves coloured by whether antibody levels declined between samples, with estimated median and 95% CrI threshold indicating 50% protection from seroconversion.

Following the first wave, to explore whether naturally induced spike-IgG prevented increases in S-IgG (a proxy for variant infection) in subsequent waves, we analysed the changes in IgG following waves 2, 3 and 4 in seropositive mothers (Table 2). For mothers seropositive after wave 1, only 28·2% (49/174) increased S-ancestral IgG after wave 2 (compared to 52% of seronegative mothers). Following wave 3, 59% of seropositive mothers responded to Delta (104/176) compared to 67% of seronegatives and following wave 4, 80% responded to Omicron (130/163) compared to 77% of seronegatives. Higher pre-wave antibody levels were associated with a lower probability of increased IgG following the subsequent wave (Table 2 and Figure 1), indicating a potentially protective effect against infection. This was more pronounced for ancestral strain antibodies against the beta wave than against the delta and omicron waves.

**Table 2:**
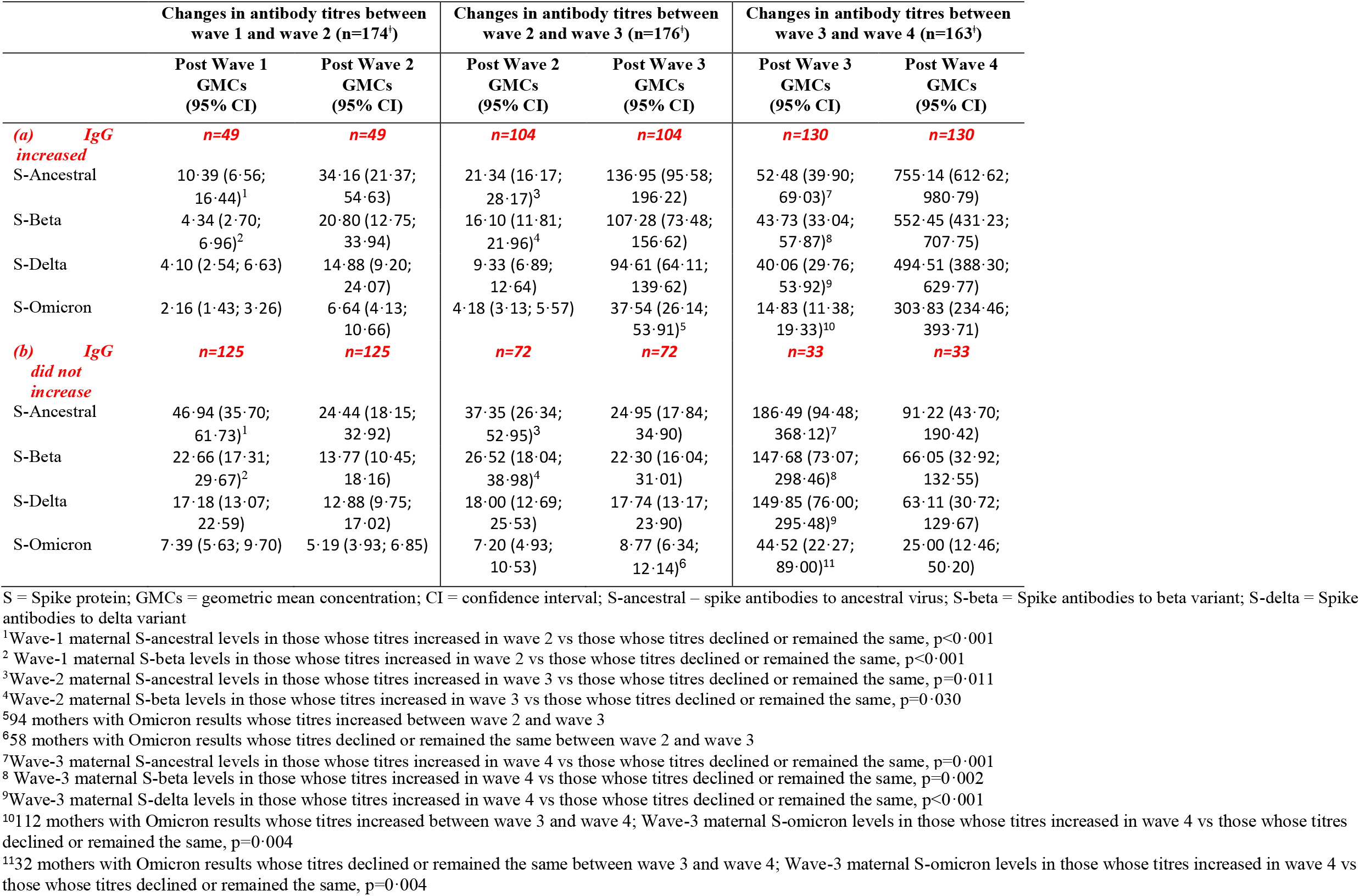
Pre and Post wave anti-spike IgG concentrations (GMC, 95% CI) in seropositive unvaccinated mothers following wave-2 (beta), wave-3 (delta) and wave-4 (omicron). Mothers have been stratified into those whose IgG increased following the wave and those whose IgG did not increase.

To explore the impact of infection induced pre-wave IgG in more detail we estimated that the probability for boosting in individuals with very low S-ancestral titres was 53% (95% CrI: 46, 64) during beta, 68% (95% CrI: 60, 85) during delta and 84% (95% CrI: 78, 90) during the omicron wave. In comparison, estimates for boosting rates in individuals with the very highest antibodies titres were 17% (95% CrI: 4, 26) 39% (95% CrI: 8, 60) and 23% (95% CrI: 2, 60) respectively (Table 3).

**Table 3.** Estimated levels of protection for minimal and maximal pre-wave WT antibody titres, 50% protection against infection (seroconversion) antibody titre threshold, and proportion of individuals with pre-wave titres above threshold.

| Wave | Probability of increased titres at minimal pre-wave antibody levels (% , 95% CrI) | Probability of increased titres at maximal pre-wave antibody levels (% , 95% CrI) | 50% protection threshold (WHO BAU/ml, median, 95% CrI) | N | N increased | Doses pre- and post-wave | Proportion of seropositives with pre-wave antibody titres higher than threshold (median, 2.5% CrI, 97.5% CrI) |  |  | N in subgroup |
| --- | --- | --- | --- | --- | --- | --- | --- | --- | --- | --- |
| Beta | 53.3 (46.0, 64.1) | 16.7 (3.5, 25.5) | 11.7 (4.4, 36.0) | 337 | 135 | 0 | 70.1% (122) | 54.6% (95) | 82.2% (143) | 337 |
|  |  |  |  |  |  | Total | 69.9% (123) | 54.5% (96) | 82.4% (145) | 339 |
| Delta | 67.7 (59.5, 85.3) | 38.9 (8.2, 60.0) | 63.6 (3.7, 757.5) | 242 | 148 | 0 | 27.7% (48) | 0.6% (1) | 89.6% (155) | 241 |
|  |  |  |  |  |  | 1 | 100.0% (1) | 0.0% (0) | 100.0% (1) | 1 |
|  |  |  |  |  |  | Total | 27.8% (70) | 0.4% (1) | 90.1% (227) | 339 |
| Omicron (threshold excluding vaccinated) | 84.3 (78.0, 90.1) | 22.5 (1.7, 58.0) | 868.5 (366.2, 2162.9) | 185 | 147 | 0 | 8.0% (13) | 2.5% (4) | 16.6% (27) | 185 |
|  |  |  |  |  |  | 1 | 65.5% (38) | 39.7% (23) | 75.9% (44) | 59 |
|  |  |  |  |  |  | 2 | 62.5% (10) | 50.0% (8) | 75.0% (12) | 16 |
|  |  |  |  |  |  | Total | 25.2% (79) | 14.7% (46) | 33.9% (106) | 339 |
| Omicron (threshold including vaccinated) | 84.3 (78.7, 89.5) | 12.0 (4.1, 23.8) | 668.6 (468.6, 920.3) | 260 | 171 | 0 | 11.0% (18) | 6.7% (11) | 14.1% (23) | 185 |
|  |  |  |  |  |  | 1 | 69.0% (40) | 62.1% (36) | 72.4% (42) | 59 |
|  |  |  |  |  |  | 2 | 62.5% (10) | 62.5% (10) | 75.0% (12) | 16 |
|  |  |  |  |  |  | Total | 27.8% (87) | 24.0% (75) | 31.0% (97) | 339 |

CrI = credible interval

Substantially greater pre-wave S-ancestral IgG titres were required to provide 50% protection against infection (a threshold defined as the inflection point of the 5-parameter logistic function) before the omicron wave, compared to the delta and beta waves (Figure 1C, 1D, 1E; Table 3). Based on these thresholds 70% (95% CrI: 55, 89), 28% (95% CrI: 1%, 90%) and 8% (95% CrI: 3, 17) of seropositive, unvaccinated participants had sufficient pre wave antibodies to be protected against infection in the beta, delta, and omicron waves respectively, table 3. These findings were also robust to the use of variant-specific titres despite a lower estimated threshold for Omicron (Table S3) as the WT and variant concentrations were highly correlated (Figure S2).

Of 154 vaccinated participants, 135 (87·7%) were seropositive prior to vaccination (Table S4). As the majority received BNT162b2 vaccine (127, 82·5%), GMCs for the BNT162b2 vaccine recipients, stratified by preceding serostatus were calculated (Figure 2 and Table S5). Antibody levels following one or two doses of vaccine were significantly higher in those who were seropositive prior to vaccine, compared to those seronegative, for all antibodies measured including S-ancestral, S-beta, S-delta and S-omicron. A 2^nd^ dose of vaccine in those seropositive prior to vaccination showed no significant increase in IgG following dose 2. In contrast a 2^nd^ dose in seronegative vaccine recipients resulted in an expected increase (Figure 2 and Table S5). A single dose of vaccine in seropositive individuals induced a higher IgG concentration than two doses in naïve vaccine recipients.

**Figure 2.**
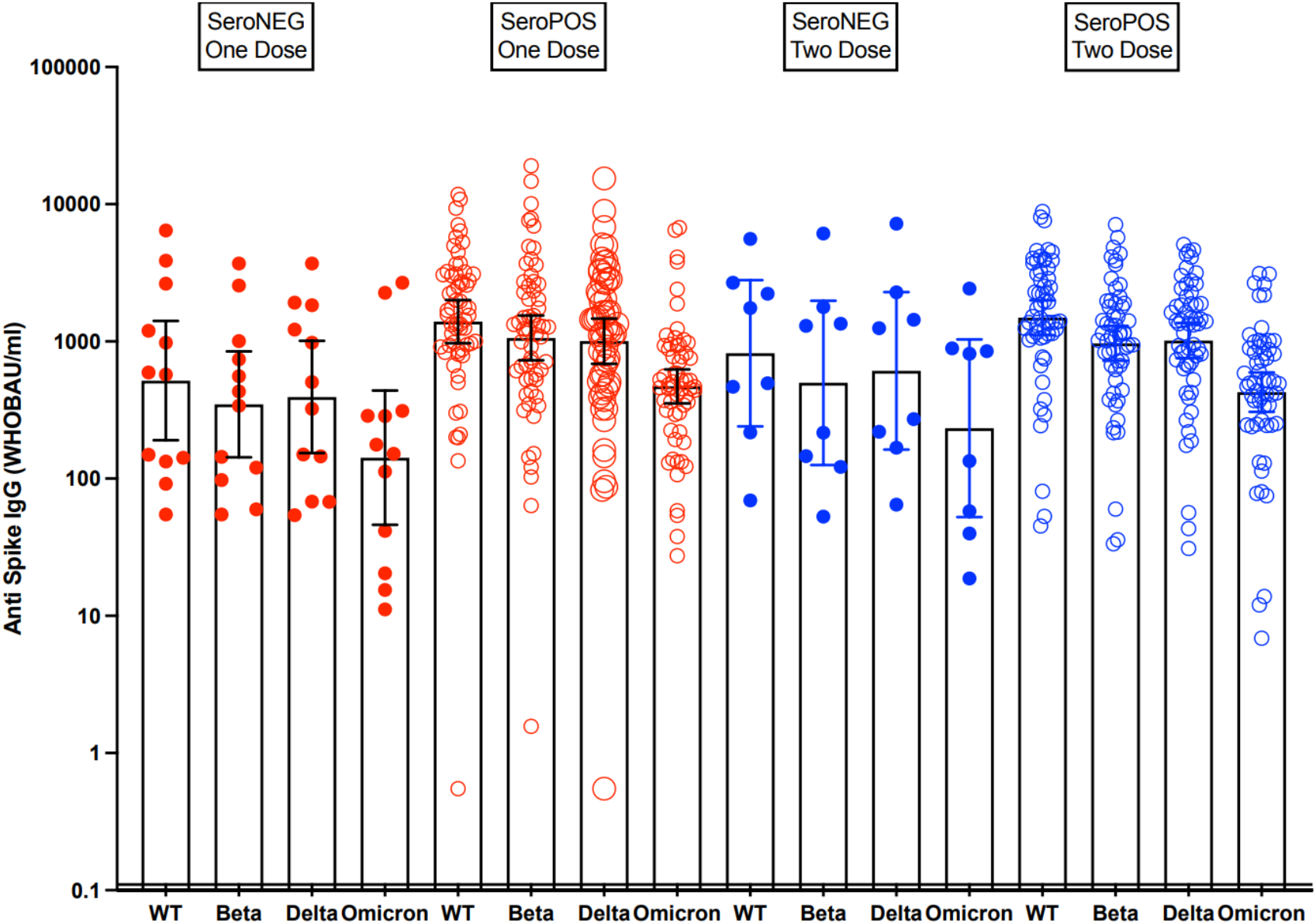
Anti-Spike IgG responses in participants after one or 2 doses of BN162b2 vaccine, stratified by serostatus prior to vaccination. One dose responses are shown in red and two doses in blue. Concentrations of seronegative vaccinees are illustrated with closed circles and seropositive with open circles.

A substantially greater proportion of seropositive (prior to vaccination), vaccinated individuals (1 and 2 doses) were above the 50% protection against infection level compared to seropositive but unvaccinated individuals in the omicron wave, with 69% (95% CrI: 62, 72) protected having received 1 dose and 63% (95% CrI: 63, 75) protected having received 2 doses, compared to 11% (95% CrI: 7, 14) protected having received 0 doses (Table 3). The estimated threshold and proportion of individuals protected were not substantially different if vaccinated individuals were excluded when calculating the threshold, indicating that antibody levels are the primary determinant of infection prevention rather than another vaccine-specific factor (50% protection threshold: 869 WHO BAU/ml (95% CrI: 366, 2163); 66% (95% CrI: 40, 76%) protected for one dose, 63% (95% CrI: 50, 75) protected for 2 doses, and 8% (95% CrI: 3, 17) protected for unvaccinated).

## Discussion

More than 2 years since the pandemic started many communities around the world have been exposed to successive waves of SARS-CoV-2 infections. This exposure has altered their susceptibility to subsequent infection^23^ and is likely responsible for the different disease profiles witnessed following the omicron wave. In communities with previous widespread exposure and vaccinations, omicron infection has been relatively mild while in communities where zero-tolerance of COVID has been pursued and thus relatively little disease-modifying population immunity has been acquired the impact of omicron has been more severe.^24^ In healthy mothers resident in the poor peri-urban area of South Africa which was the focus of this study, 53% were seropositive after the first wave of ancestral SARS-CoV-2 and this rose, in unvaccinated mothers, to 97·3% following the omicron wave which swept South Africa between November 2021 and January 2022. This rate of seropositivity in an unvaccinated population is to our knowledge the highest reported and greatly exceeding population estimates for Africa of 65·7%.^25^ Despite high rates of successive exposure to SARS-CoV-2 very few of the mothers in this study tested positive for SARS-CoV-2 (5·3%), despite ready access to health care facilities, and only 3 were hospitalized. This concurs with the recently published WHO analysis suggesting Africa differentiates itself from other regions by its high number of asymptomatic (67%) infections^25^ as well as a South African based household infection study which estimated that 85·3% of infections were asymptomatic.^26^

Prior exposure resulting in an immune response to SARS-CoV-2 was associated with a reduced likelihood of infection.. This finding is consistent with a reduced risk of reinfection in a household study conducted in Soweto where prior infection provided durable protection against reinfection throughout the study period which included the beta and delta waves.^27^ Despite antibody levels being in the range of or exceeding those following the beta wave, a substantially lower proportion of seropositive individuals had pre-wave antibody levels above the modelled threshold of protection in the delta and omicron waves (7% and 4% respectively vs. 44% for beta), indicating lower cross-protection of antibodies against these variants. Rates of infection in the seronegative and seropositive mothers following the omicron wave were similar and only those with very high natural antibodies had a reduced risk of infection explaining the omicron variants propensity for high rates of both primary and reinfection.^5^

In seropositive mothers, responses to BNT162b2 were higher after one or two doses, compared to the seronegative mothers. As a consequence of higher titres, a greater proportion of vaccinated mothers had antibody levels above the 50% threshold providing protection from omicron infection compared to mothers with naturally acquired immunity. While 2 doses of COVID-19 vaccines fail to provide durable protection against omicron infections, booster doses have been associated with good short term effectiveness against omicron infection.^14,28,29^ This prevention of infection is likely related to the higher concentrations of cross reactive IgG induced by the booster.

Hybrid immunity, that seen after a combination of natural and vaccine induced immunity, is becoming important as an increasing proportion of the global population becomes exposed to SARS-CoV-2 and more vaccination takes place on the background of natural immunity. In our study, responses to either one or two doses of BNT162b2 in seropositive individuals were greater than those seen in naïve vaccinees and a second dose in a naturally infected individual was not associated with the expected increase in IgG. It is also now well recognized that vaccination following natural exposure is associated with great vaccine effectiveness^13-15^ compared to either natural or vaccine induced immunity alone. As suggested by our study the greater effectiveness of hybrid immunity is likely linked to enhanced immunogenicity of vaccine on the background of natural immunity and the reported increase in the breadth of immunity.^18^ We were also able to demonstrate qualitative differences following exposure to variants between naïve and seropositive mothers with naïve mothers mounting an IgG response dominated by the spike antigen from the VOC while seropositive mothers responded with a dominant wild type IgG irrespective of the VOC they were exposed to suggesting a degree of imprinting as first suggested by Röltgen and colleagues.^18^ As there are important differences in the immunogenicity and effectiveness of vaccines when administered to previously exposed individuals, the need to provide 2 doses of vaccine to unvaccinated individuals as a priming schedule should be reconsidered. With high (and increasing) rates of seropositivity in many unvaccinated communities in the world, there may be a substantial advantage in focusing efforts on providing a single dose of vaccine to such communities rather than having targets for two dose priming. It is possible that two doses in a seropositive individual will provide more durable immunity^13^ but the relative benefits of a second dose require further study.

Our study has several limitations including that infection to a variant was inferred from an increase in anti-spike IgG. As individuals were not tested for active infection unless symptomatic, we were unable to determine whether individuals were exposed during the course of a wave unless seroconversion occurred; hence, those who did not seroconvert during a wave may contain a mixture of those who were exposed and experienced an aborted infection due to sterilizing immunity, and those who were unexposed. Additionally a degree of antibody waning may have taken place between pre-wave sampling and exposure in the subsequent wave, so antibody levels at exposure are likely to have been lower than when measured. We did not measure neutralizing antibody as we have previously shown excellent correlation between binding antibody as measured in our laboratory and live virus or pseudovirus neutralization.^12^ We also did not have access to stored cells to evaluate cellular immune mechanism although these may be more important for disease/serious disease manifestation rather than the prevention of infection.

In summary this study has shown very high seroprevalence to SARS-CoV-2 in a poor, peri-urban South African community. Seropositivity via natural exposure to SARS-CoV-2 was associated with subsequent protection from infection with beta and delta variant but not Omicron, where only very high levels of natural antibody provided protection. Vaccination of seropositive individuals elicited higher concentrations of Spike IgG compared to seronegative mothers and a greater proportion of seropositive vaccinated mothers were therefore protected from Omicron. A single dose of current vaccine based on Wild Type SARS-CoV-2 in seropositive individuals may provide sufficient protection against known or related SARS-CoV-2 variants.

## Data Availability

An anonymised, de-identified version of the dataset can be made available upon request to
allow all results to be reproduced.

## Author contributions

HJZ contributed to conceptualisation, funding acquisition, methodology, supervision and writing of original draft. RM, LW did data curation and formal analysis. MB, TB were involved in methodology and project administration. MJ, AH contributed to laboratory investigation and methodology. MPN was involved in conceptualisation and methodology. BJQ, SF undertook formal analysis. DG contributed to conceptualisation, formal analysis, methodology, supervision and writing of original draft. All authors contributed to the final manuscript.

## Data sharing

An anonymised, de-identified version of the dataset can be made available upon request to allow all results to be reproduced.

## Acknowledgements

We thank the children and families participating in the DCHS. We acknowledge the study staff, and the clinical and administrative staff of the Western Cape Government Health Department for their support of the study and Jim Wilbur from Meso Scale Discovery for technical support. This study was funded by the UK NIHR GECO award (GEC111), the Wellcome Trust Centre for Infectious Disease Research in Africa (CIDRI), the Bill & Melinda Gates Foundation, USA (grant number OPP1017641, OPP1017579) and the NIH H3 Africa (grant numbers U54HG009824, U01AI110466]. HZ is supported by the SA-MRC. MPN is supported by an Australian National Health and Medical Research Council Investigator Grant (APP1174455). BJQ is supported by a grant from the Bill and Melinda Gates Foundation (OPP1139859). Stefan Flasche is supported by a Sir Henry Dale Fellowship jointly funded by the Wellcome Trust and the Royal Society (Grant number 208812/Z/17/Z). DG benefits from the support of the NIHR GOSH Biomedical Research Centre.

## Supplementary Materials

[DCHS Supplementary material 0706 FINAL formatted.docx](https://www.editorialmanager.com/thelancetid/download.aspx?id=488956&guid=d90275d5-c3bc-47f0-8eb5-09db291999f0&scheme=1)

