## Supplementary material for "Natural and hybrid immunity following four COVID-19 waves in a South African cohort": Table S1, Table S2, Table S3, Table S4, Table S5, Figure S1, Figure S2

**Table S1. Characteristics of participants by seroprevalence in first wave**

|  | **All** | **SARS-CoV2-S seropositive** | **SARS-CoV2-S seronegative** | **p-value** |
| --- | --- | --- | --- | --- |
| **Maternal characteristics** | n=339 | n=176 (51·9%) | n=163 (48·1%) |  |
| Age [median (IQR)] years | 32·9 (28·9; 37·2) | 33·5 (28·9; 38·3) | 32·1 (28·9; 36·8) | 0·082 |
| HIV infected, n (%) | 69 (20·4) | **45 (25·6)** | **24 (14·7)** | **0·013** |
| Smoker, n (%) | 124 (36·6) | **50 (28·4)** | **74 (45·4)** | **<0·001** |
| Asthma/COPD, n(%) | 10 (3·0) | 5 (2·8) | 5 (3·1) | 0·902 |
| Diabetes, n(%) | 2 (0·6) | 1 (0·6) | 1 (0·6) | 0·731 |
| Weight [median (IQR)] | 72·6 (59·1; 89·9) | 72·6 (62·0; 91·5) | 72·8 (55·5; 88·8) | 0·075 |
| **Socio-demographic characteristics** | | |  |  |
| Marital status – single, n(%) | 188 (55·5) | 92 (52·2) | 96 (58·9) | 0·220 |
| Married/cohabiting, n(%) | 151 (44·5) | 84 (47·7) | 67 (41·1) |  |
| Maternal education, n(%) |  |  |  |  |
| Primary education | 21 (6·2) | 9 (5·1) | 12 (7·4) | 0·579 |
| Some secondary education | 190 (56·1) | 95 (54·0) | 95 (58·3) |  |
| Completed secondary education | 115 (33·9) | 65 (36·9) | 50 (30·7) |  |
| Tertiary education, n(%) | 13 (3·8) | 7 (4·0) | 6 (3·6) |  |
| Maternal employment, n(%) | 131 (38·6) | 72 (40·9) | 59 (36·2) | 0·373 |
| Household income per month, n(%) |  |  |  |  |
| <1000 ZAR (<60 USD) | 56 (16·5) | 25 (14·2) | 31 (19·0) | 0·345 |
| 1000-5000 ZAR (60-300 USD) | 221 (65·2) | 115 (65·3) | 106 (65·0) |  |
| >5000 ZAR (>300 USD) | 62 (18·3) | 36 (20·5) | 26 (16·0) |  |
| Number of household members [median (IQR)] | 5 (4; 6) | 5 (4; 6) | 5 (4; 6) | 0·282 |

Abbreviations: CoV2-S spike, IQR=interquartile range; ZAR= South African Rand; USD= United States dollar

**Table S2. Multivariate analysis of factors associated with seropositivity^*^**

|  | **Unadjusted OR (95% CI)** | **Adjusted OR (95% CI)**** |
| --- | --- | --- |
| Wave | **3·06 (2·61; 3·59)** | **3·22 (2·71; 3·82)** |
| Age | **1·05 (1·02; 1·08)** | 1·02 (0·98; 1·06) |
| HIV infection | **1·68 (1·06; 2·66)** | 1·63 (0·92; 2·89) |
| Current smoking | **0·50 (0·36; 0·70)** | **0·43 (0·28; 0·66)** |
| Asthma | 0·78 (0·33; 1·85) | 0·77 (0·28; 2·15) |
| Weight | **1·01 (1·00; 1·02)** | 1·00 (0·99; 1·01) |
| Marital status – single | Reference | Reference |
| Married/cohabiting | 1·06 (0·76; 1·49) | 1·03 (0·67; 1·59) |
| Maternal education |  |  |
| Primary education | Reference | Reference |
| Some secondary education | 1·05 (0·53; 2·07) | 1·30 (0·58; 2·94) |
| Completed secondary education | 1·16 (0·57; 2·35) | 1·58 (0·66; 3·76) |
| Tertiary education | 0·89 (0·32; 2·25) | 1·05 (0·26; 4·24) |
| Maternal employment | 1·19 (0·84; 1·68) | 0·93 (0·59; 1·46) |
| Household income per month |  |  |
| <1000 ZAR (<60 USD) | Reference | Reference |
| 1000-5000 ZAR (60-300 USD) | 1·48 (0·96; 2·28) | 1·05 (0·57; 1·93) |
| >5000 ZAR (>300USD) | 1·60 (0·92; 2·80) | 1·61 (0·76; 3·43) |
| Household size | 1·08 (0·99; 1·18) | **1·14 (1·02; 1·27)** |

OR=Odds ratio; CI=confidence interval

*Seropositive defined as S-antibodies to ancestral virus ≥ 1·09 WHO BAU/ml

**339 women included, with 1105 observations; observations after vaccination excluded from analysis

| Variant IgG assessed | Probability of increased titres at minimal pre-wave antibody levels (%, 95% CrI) | Probability of increased titres at maximal pre-wave antibody levels (%, 95% CrI) | 50% reduction threshold (WHO BAU/ml, median, 95% CrI) | N | N increased | Doses pre and post-wave | Proportion of seropositives with pre-wave antibody titres higher than threshold (median) | Proportion of seropositives with pre-wave antibody titres higher than threshold (2·5% CrI) | Proportion of seropositives with pre-wave antibody titres higher than threshold (97·5% CrI) |
| --- | --- | --- | --- | --- | --- | --- | --- | --- | --- |
| Beta | 51·4 (43·2, 77·2) | 19·9 (1·6, 30·7) | 6·3 (1·3, 31·0) | 337 | 134 | 0 | 75·8% (119) | 42·0% (66) | 98·7% (155) |
|  |  |  |  |  |  | Total | 75·5% (120) | 42·1% (67) | 98·7% (157) |
| Delta | 76·2 (69·2, 82·8) | 42·3 (19·9, 63·9) | 36·9 (15·3, 159·5) | 232 | 164 | 0 | 23·0% (35) | 4·6% (7) | 49·3% (75) |
|  |  |  |  |  |  | Total | 24·2% (54) | 3·6% (8) | 52·5% (117) |
| Omicron | 86·3 (79·5, 92·0) | 13·1 (3·7, 25·7) | 185·0 (117·2, 278·6) | 217 | 142 | 0 | 10·5% (15) | 5·6% (8) | 14·7% (21) |
|  |  |  |  |  |  | 1 | 69·1% (38) | 61·8% (34) | 74·5% (41) |
|  |  |  |  |  |  | 2 | 62·5% (10) | 62·5% (10) | 68·8% (11) |
|  |  |  |  |  |  | Total | 28·0% (79) | 24·1% (68) | 32·6% (92) |

**Table S3· Estimated levels of protection for minimal and maximal pre-wave variant-specific antibody titres, 50% protection against seroconversion antibody titre threshold, and proportion of individuals with pre-wave titres above threshold.**

**Table S4· Vaccinations and serostatus prior to vaccine**

|  | **number of vaccines (n=154, %)** | **Seropositive prior to vaccine (n=135, %)** | **Median days between vaccine and blood sample (IQR)** |
| --- | --- | --- | --- |
| **Vaccine** |  |  |  |
| Ad26.COV.2.S | 27 (17·4) | 26 (19·3) | 64 (33; 113) |
| BNT162b2– 1 dose only | 61 (39·6) | 51 (37·8) | 32 (14; 56) |
| BNT162b2– 2 doses | 66 (42·6) | 58 (43·0) | 75 (39; 97·5) |

IQR=Interquartile range

**Table S5 GMCs of participants after 1 or 2 doses of BNT162b2 (Pfizer-BioNTech)** **vaccine stratified by serostatus prior to vaccination**

|  | **One dose (n=72)** | | **Two doses (n=65)** | |
| --- | --- | --- | --- | --- |
|  | **Seronegative prior to vaccine (n=12)** | **Seropositive prior to vaccine (n=60)** | **Seronegative prior to vaccine (n=8)** | **Seropositive prior to vaccine (n=57)** |
|  | **GMC (95% CI)** | **GMC (95% CI)** | **GMC (95% CI)** | **GMC (95% CI)** |
| S-ancestral | 517·92 (190·41; 1408·79)^1^ | 1590·14 (1231·33; 2053·50)^1^ | 819·71 (239·97; 2800·00)^5^ | 1488·78 (1109·50; 1997·70)^5^ |
| S-beta | 348·22 (143·05; 847·67·25)^2^ | 1181·93 (865·85; 1613·38)^2^ | 499·16 (125·69; 1982·26)^6^ | 967·92 (721·63; 1298·28)^6^ |
| S-delta | 393·38 (153·45; 1008·46)^3^ | 1139·07 (852·57; 1521·84)^3^ | 609·91 (162·73; 2286·00)^7^ | 1012·06 (753·54; 1359·28)^7^ |
| S-omicron | 142·15 (46·01; 439·20)^4^ | 469·98 (353·07; 625·60)^4^ | 233·02 (52·50; 1034·25)^8^ | 427·42 (307·18; 594·72)^8^ |

GMC= Geometric mean concentration; CI= confidence interval

^1^ Comparison of GMC S-ancestral titres of 1 dose in seropositive vs seronegative participants, p=0·018

^2^ Comparison of GMC S-beta titres of 1 dose in seropositive vs seronegative participants, p=0·008

^3^ Comparison of GMC S-delta titres of 1 dose in seropositive vs seronegative participants, p=0·026

^4^ Comparison of GMC S-omicron titres of 1 dose in seropositive vs seronegative participants, p=0·007

^5^ Comparison of GMC S-ancestral titres of 2 doses in seropositive vs seronegative participants, p=0·299

^6^ Comparison of GMC S-beta titres of 2 doses in seropositive vs seronegative participants, p=0·299

^7^ Comparison of GMC S-delta titres of 2 doses in seropositive vs seronegative participants, p=0·328

^8^ Comparison of GMC S-omicron titres of 2 doses in seropositive vs seronegative participants, p=0·485

**Figure S1. Flow chart of participants across 4 waves**

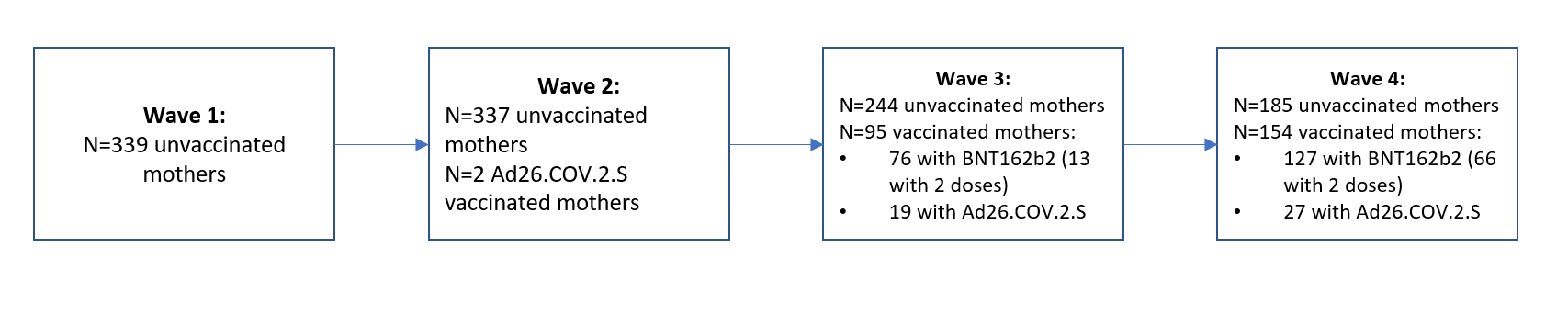

**Figure S2. Correlations of wild-type and variant titres, pre and post-wave.**

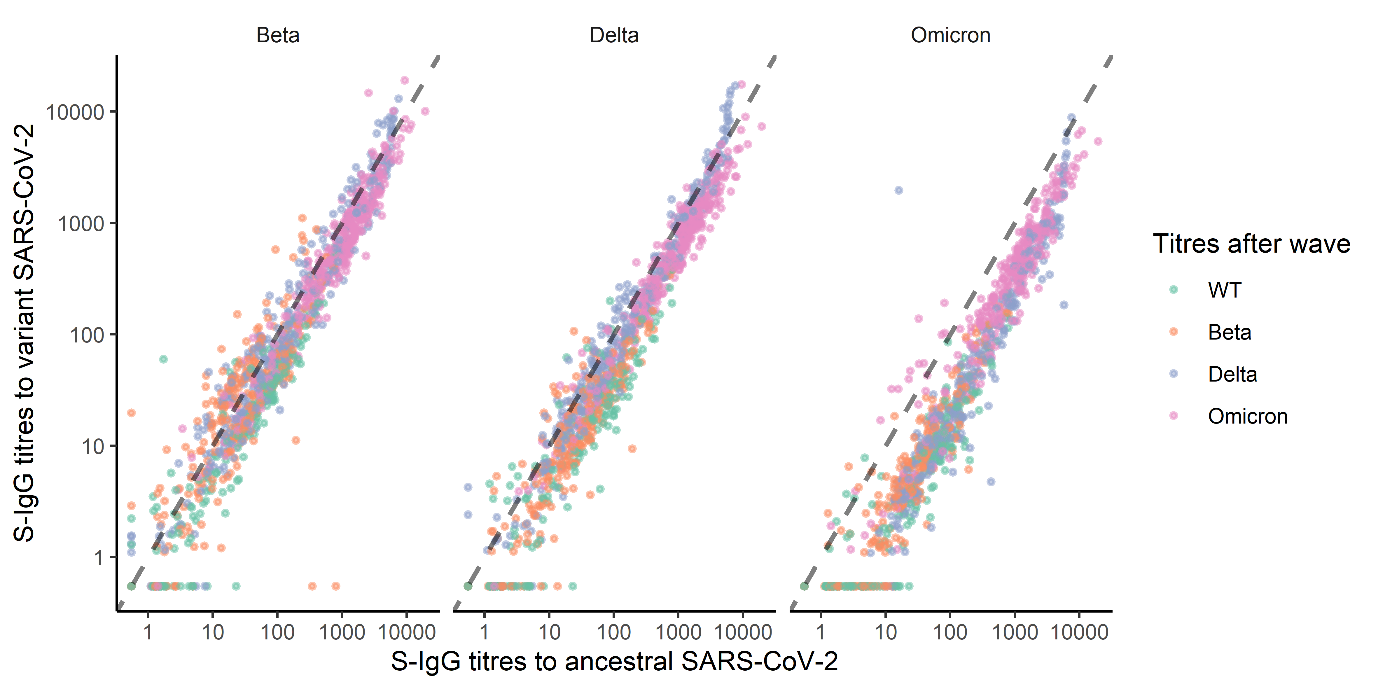
